# Disaster preparedness among women with a recent live birth in Hawaii, a cross-sectional study, results from the Pregnancy Risk Assessment Monitoring System (PRAMS), 2016

**DOI:** 10.1101/2021.04.14.21255501

**Authors:** Penelope Strid, Carlotta Ching Ting Fok, Marianne Zotti, Holly B. Shulman, Jane Awakuni, L. Duane House, Brian Morrow, Judy Kern, Matthew Shim, Sascha Ellington

**Author notes:** **Corresponding author** Penelope Strid; U.S. Centers for Disease Control and Prevention, 4770 Buford Highway NE, Mailstop S 107-2, Atlanta, GA 30341.

## Abstract

**Objectives:** This study examines emergency preparedness behaviors among women with a recent live birth in Hawaii.

**Methods:** Using the 2016 Hawaii Pregnancy Risk Assessment Monitoring Survey we estimated weighted prevalence of eight preparedness behaviors.

**Results:** Among 1010 respondents (weighted response rate=56.3%), 79.3% reported at least one preparedness behavior and 11.2% performed all eight behaviors. The prevalence of women with a recent live birth in Hawaii reporting preparedness behaviors includes: 63.0% (95% CI: 58.7-67.1%) having enough supplies at home for at least seven days, 41.3% (95% CI: 37.1-45.6%) having an evacuation plan for their child(ren), 38.7% (95% CI: 34.5, 43.0) having methods to keep in touch, 37.8% (95% CI: 33.7, 42.1) having an emergency meeting place, 36.6% (95% CI: 32.6, 40.9) having an evacuation plan to leave home, 34.9% (95% CI: 30.9, 39.2) having emergencies supplies to take with if they have to leave quickly, 31.8% (95% CI: 27.9, 36.0) having copies of important documents, 31.6% (95% CI: 27.7, 35.8) having practiced what to do.

**Conclusion:** One in ten women practiced all eight behaviors indicating more awareness efforts are needed among this at-risk population in Hawaii. Hawaii can measure the effect of interventions to increase preparedness by tracking this question over time.

**Significance:** *“What is already known on this subject?”:* Preparedness is associated with reduced vulnerability, and postpartum women are considered an at-risk population in the post-disaster period with special clinical needs. One prior study has assessed disaster preparedness among postpartum women.

*“What this study adds?”:* This is the first study to describe a methodology to analyze the eight-part PRAMS emergency preparedness question. Among recently postpartum women in Hawaii, about 80% practiced at least one of eight emergency preparedness measures assessed and about 10% practiced all behaviors.

## Introduction

From 2000 to 2016, the United States (US) averaged 58 Federal Emergency Management Agency (FEMA) defined major disaster declarations annually (Federal Emergency Management Agency 2020). During this same period, Hawaii (HI) experienced 11 FEMA major disaster declarations (Federal Emergency Management Agency 2020). In the past 20 years, major disasters have occurred more frequently in the US; reinforcing the need for disaster preparation to mitigate damage and harm (Federal Emergency Management Agency 2020; Yahmed and Koob 1996; World Health Organization 2007).

Disasters can compound and exacerbate social vulnerabilities (Morrow 1999; Nour 2011). According to the Pandemic and All-Hazards Preparedness and Advancing Innovation Act (2019), pregnant and postpartum women and infants are considered an at-risk population with special clinical needs. Basic resources such as clean water, nutritious foods, diapers, and safe sleeping areas may not be readily available but are especially important for pregnant and breastfeeding women and their infants (Callaghan et al. 2007; Ewing, Buchholtz, and Rotanz 2008; Larrance, Anastario, and Lawry 2007). Public health and medical services may be interrupted due to damaged infrastructure, power outages, and lack of trained personnel (Harville, Xiong, and Buekens 2009).

Vulnerable populations such as pregnant and postpartum women may have a slow recovery after a disaster further amplified as a result of socioeconomic disadvantages, limited financial resources, and poor social support (Morrow 1999; Nour 2011; Zahran, Peek, Snodgrass, Weiler, and Hempel 2011; Callaghan et al. 2007; Giarratano, Barcelona, Savage, and Harville 2019). Preparedness is associated with reduced vulnerability (Yahmed and Koob 1996; World Health Organization 2007). Baseline measures of preparedness can inform public health education campaigns to increase preparedness and help in planning to meet the population’s needs during a disaster. Limited information is available on the prevalence of preparedness among pregnant and postpartum women (Zilversmit, Sappenfield, Zotti, and McGehee 2014; Maher 2019; National Institutes of Health 2019; Hapsari, Jayanti, Nugraheni, and Panuntun 2020). It may be difficult to estimate baseline measures of preparedness among pregnant and postpartum women through traditional population-based sampling since they are a small percentage of the general population (Horney, Zotti, Williams, and Hsia 2012). The Pregnancy Risk Assessment Monitoring System (PRAMS) is an annual survey conducted by states and the Centers for Disease Control and Prevention (CDC) which can be used to estimate baseline preparedness among women who recently had a live birth. An analysis of one 2009 Arkansas PRAMS emergency preparedness question provided the first prevalence estimate of postpartum women who responded that they had an emergency preparedness plan (Zilversmit, Sappenfield, Zotti, and McGehee 2014). In 2016, PRAMS introduced an eight-part supplemental question to assess disaster preparedness (Centers for Disease Control and Prevention 2016). Using the 2016 Hawaii PRAMS data, we assessed the prevalence of emergency preparedness behaviors among women that recently had a live birth. To look at characteristics associated with preparedness, we used factor analysis to distill the eight preparedness behaviors into correlated preparedness factors.

## METHODS

### Data Collection

PRAMS is a state, population-based sample of women who recently had a live birth. State birth certificate files are used to select approximately 200 women each month in Hawaii. The surveys are self-reported and follow the systematic PRAMS methodology; previously described by Shulman et al. (2018). Women receive the survey approximately two months after delivery. Contact is made initially by mail and then by phone. In Hawaii, PRAMS is only offered in English. Those who participate receive a $10 gift card to a local food market. Consent by mail is implied by returning a completed questionnaire; verbal consent is provided by phone. The protocol is IRB reviewed and approved by the CDC and the Hawaii State Department of Health.

### Variables

The eight pre-tested, standardized PRAMS emergency preparedness questions are listed in Table 1 (Centers for Disease Control and Prevention 2016). Data from the Hawaii birth certificate included maternal age, maternal education, marital status, urban or rural residence, maternal race, and participation in Special Supplemental Nutrition Program for Women Infants and Children (WIC) during pregnancy. Age was collapsed into five categories: 19 years or less, 20 to 24 years, 25 to 29 years, 30 to 34 years, and 35 years or greater. Education was categorized as less than high school diploma, high school diploma or equivalent, some college, and baccalaureate degree or higher. The urban or rural residence variable was established by county, and Maui was classified as rural according to the 2012 Census classification. Race was categorized by the racial categories specific to Hawaiian birth certificates: White, Native Hawaiian, Filipino, Japanese, Other Pacific Islander, and other. Anyone selecting Native Hawaiian or part-Hawaiian was grouped into the Native Hawaiian race. Other Pacific Islanders included those selecting the category, and Samoans and Guamanians. All other races were categorized as “other”, consistent with methods described by Sorenson et al. (2003).

**Table 1.**
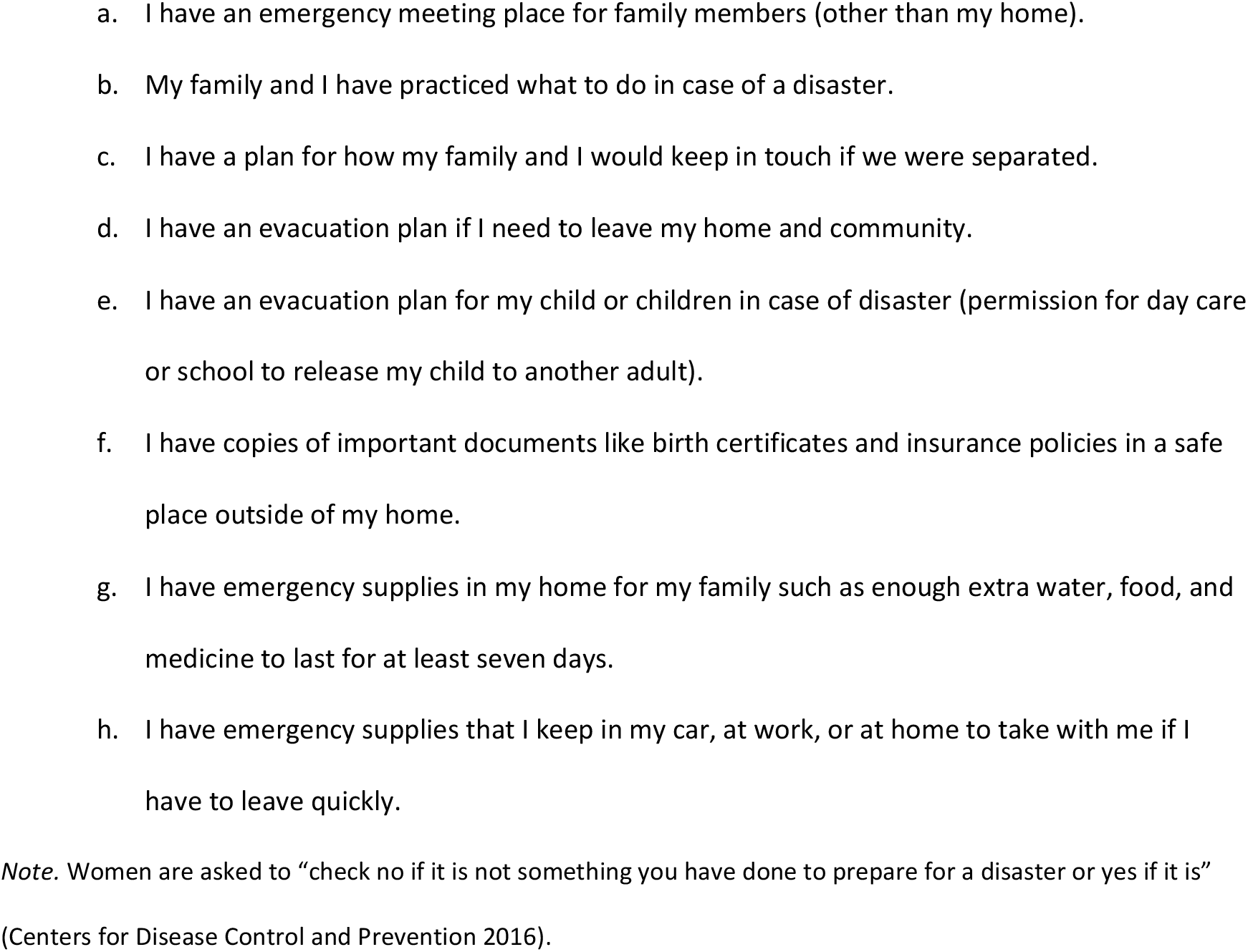
Eight standardized emergency preparedness questions from 2016 Hawaii PRAMS

Information obtained from PRAMS included current health insurance at time of survey, total income, and family size. Insurance status was categorized into three options: private, public, or none. Individuals that indicated having both private and public insurance were categorized as having private insurance. Military insurance was captured as a separate option on the survey and categorized as private for the analysis. Total income captured the reported household income 12 months prior to the birth, and family size was defined as number of individuals living on the total income in the 12 months prior to the live birth. Family sizes of five or greater were collapsed into a single category. A variable categorizing income as a percent of the federal poverty level (FPL) was developed from total income and family size, compared to the January 2016 poverty guidelines for Hawaii issued by the Federal Register of the Department of Health and Human Services (Department of Health and Human Services 2016).

Four FPL groups are reported as less than or equal to 100%, 101-185%, 186-300%, and greater than 300% FPL, consistent with federal programing eligibility and Hawaii PRAMS trend report (Hawaii 2019; Department of Health and Human Services 2016).

### Statistical Analysis

The sample is weighted to be representative of the state by accounting for sampling stratification, nonresponse, and noncoverage. The sample is stratified initially by county (Honolulu, Maui, Hawaii, and Kauai) and within Honolulu county, by birthweight.

Factor analysis was performed to assess underlying factors among the eight emergency preparedness behaviors. The dataset was converted to a tetrachoric correlation matrix, and factor analysis was performed using the maximum likelihood estimate and varimax rotation. The overall measure of sampling adequacy was 0.91 suggesting factor analysis could be used for these data (Kaiser 1974).

Data analysis was performed in SAS-callable SUDAAN 11.0 (SAS v9.4) to account for weighted data and complex survey methods. Multivariable logistic regression models were used to estimate predicted marginal prevalence ratios to assess characteristics of women associated with emergency preparedness behaviors. White was the referent category for race, otherwise the subcategory with the greatest proportion of women was used as the referent category. Variables included in the model were established *a priori* from existing literature and included age, education, marital status, urban or rural residence, race, insurance status, family size, FPL, and use of WIC during pregnancy.

## RESULTS

### Study Characteristics

In 2016, 1,999 women were contacted for PRAMS in Hawaii and 1,076 responded (weighted response rate 56.3%). The final sample for this analysis included 1,010 women; 66 were excluded for not answering any of the emergency preparedness questions. Table 2 presents the demographic characteristics of the 1,010 women included in this analysis.

**Table 2.** Demographic characteristics of women with a recent live birth in Hawaii, 2016 Hawaii PRAMS (N=1,010)

|  | N | (%) | 95% CI |
| --- | --- | --- | --- |
| **Age (years)** |  |  |  |
| ≤19 | 33 | (3.0) | (1.9, 4.8) |
| 20-24 | 156 | (14.5) | (11.7, 17.9) |
| 25-29 | 288 | (28.5) | (24.8, 32.6) |
| 30-34 | 313 | (31.2) | (27.3, 35.3) |
| ≥35 | 220 | (22.8) | (19.3, 26.6) |
| **Education<sup>a</sup>** |  |  |  |
| Less than High School | 147 | (17.1) | (13.8, 20.9) |
| High School | 174 | (18.9) | (15.8, 22.5) |
| Some College | 336 | (29.3) | (25.6, 33.3) |
| Baccalaureate or Higher | 352 | (34.7) | (30.7, 38.9) |
| **Marital status** |  |  |  |
| Married | 604 | (63.4) | (59.2, 67.4) |
| Not Married | 406 | (36.6) | (32.6, 40.8) |
| **Residence** |  |  |  |
| Rural | 590 | (27.7) | (26.9, 28.5) |
| Urban | 420 | (72.3) | (71.5, 73.1) |
| **Race<sup>b</sup>** |  |  |  |
| White<sup>c</sup> | 227 | (21.6) | (18.2, 25.3) |
| Native Hawaiian<sup>d</sup> | 314 | (30.3) | (26.5, 34.4) |
| Filipino | 195 | (16.9) | (13.9, 20.3) |
| Japanese | 102 | (12.1) | (9.5, 15.3) |
| Other Pacific Islander<sup>e</sup> | 50 | (5.7) | (4.0, 8.1) |
| Other<sup>f</sup> | 119 | (13.5) | (10.7, 16.9) |
| **Insurance<sup>g</sup>** |  |  |  |
| Private<sup>h</sup> | 624 | (69.1) | (65.1, 72.8) |
| Public | 332 | (27.2) | (23.7, 31.1) |
| None | 41 | (3.7) | (2.5, 5.7) |
| **Family size<sup>i,j</sup>** |  |  |  |
| 1 | 113 | (10.2) | (7.9, 13.1) |
| 2 | 310 | (30.4) | (26.5, 34.5) |
| 3 | 249 | (28.5) | (24.6, 32.6) |
| 4 | 194 | (18.6) | (15.5, 22.3) |
| ≥5 | 120 | (12.4) | (9.7, 15.6) |
| **% FPL<sup>k</sup>** |  |  |  |
| ≤100% | 252 | (24.3) | (20.8, 28.3) |
| 101-185% | 254 | (25.3) | (21.6, 29.3) |
| 186-300% | 167 | (17.2) | (14.1, 20.8) |
| >300% | 273 | (33.2) | (29.1, 37.6) |
| **WIC during pregnancy<sup>l</sup>** |  |  |  |
| Yes | 337 | (35.0) | (30.9, 39.4) |
| No | 609 | (65.0) | (60.6, 69.1) |

Note. Reported sample sizes are unweighted and reported percentages are weighted. Abbreviations: Federal Poverty Level (FPL); Special Supplemental Nutrition Program for Women, Infants, and Children (WIC) <sup>a</sup>N=1,009. <sup>b</sup>N=1,008. <sup>c</sup>Includes Portuguese. <sup>d</sup>Includes mixed race Native Hawaiians. <sup>e</sup>Includes Samoan, Guamanian, Other Pacific Islander. <sup>f</sup>Includes American Indian, Alaska Native, Asian Indian, Black, Chinese, Cuban, Mexican, Mixed Race, other Asian, Puerto Rican, and other. <sup>g</sup>N=997. <sup>h</sup>Includes military healthcare. <sup>i</sup>N=986. <sup>j</sup>Family size describes number of individuals living on total income in the 12 months prior to the birth. <sup>k</sup>Percent FPL determined by reported household income and number of individuals living on that income 12 months prior to birth, compared to the January 2016 poverty guidelines for Hawaii issued by the Federal Register of the Department of Health and Human Services. <sup>l</sup>N=946.

The largest proportion of respondents for each demographic characteristic were aged 30-34 (31.2%), had completed a baccalaureate degree or higher (34.7%), were married (63.4%), lived in an urban area (72.3%), were Native Hawaiian race (30.3%), had private insurance at the time of the survey (69.1%), had a family size of two (30.4%), had an income greater than 300% FPL (26.5%), and did not use WIC (65.0%).

### Preparedness Behaviors

Among all women, 79.3% (n=826) reported at least one preparedness behavior, and 11.2% (n=96) reported all eight. Having emergency supplies was most commonly reported with 63.0% (95% CI: 58.7-67.1%) of women having enough supplies at home for at least seven days, followed by 41.3% (95% CI: 37.1-45.6%) having an evacuation plan for their child(ren). Less than 40% of women reported other preparedness behaviors: having an emergency meeting place (37.8%, 95% CI: 33.7-42.1%); practicing what to do in case of a disaster (31.6%, 95% CI: 27.7-35.8%); having a plan to keep in touch with family if separated (38.7%, 95% CI: 34.5-43.0%); having an evacuation plan to leave home (36.6%, 95% CI: 32.6-40.9%); having copies of important documents (31.8%, 95% CI: 27.9-36.0%); and having emergency supplies to take during an evacuation (34.9%, 95% CI: 30.9-39.2%). (Table 3)

**Table 3.** Prevalence of disaster preparedness behaviors among women with a recent live birth in Hawaii, 2016 Hawaii PRAMS

|  | N | (%) | 95% CI |
| --- | --- | --- | --- |
| **Individual Questions** |  |  |  |
| a. Emergency meeting place | 373 | (37.8) | (33.7, 42.1) |
| b. Practiced what to do | 304 | (31.6) | (27.7, 35.8) |
| c. Keeping in touch | 390 | (38.7) | (34.5, 43.0) |
| d. Evacuation plan to leave home and community | 353 | (36.6) | (32.6, 40.9) |
| e. Evacuation plan for child(ren) | 407 | (41.3) | (37.1, 45.6) |
| f. Copies of important documents | 315 | (31.8) | (27.9, 36.0) |
| g. Emergency supplies in home to last for at least 7 days | 666 | (63.0) | (58.7, 67.1) |
| h. Emergency supplies to take with if had to leave quickly | 348 | (34.9) | (30.9, 39.2) |
| Factor 1 (Plans)<sup>a</sup> | 610 | (59.8) | (55.5, 64.0) |
| Factor 2 (Documents)<sup>b</sup> | 315 | (31.8) | (27.9, 36.0) |
| Factor 3 (Supplies)<sup>c</sup> | 697 | (66.8) | (62.5, 70.8) |
| No preparedness behaviors | 184 | (20.7) | (17.3, 24.6) |

*Note.* Reported sample sizes are unweighted and reported percentages are weighted.
<sup>a</sup>Answered yes to at least one behavior among: have an emergency meeting place, have practiced what to do in case of a disaster, have a plan for how family would keep in touch if separated, have an evacuation plan if she needed to leave her home and community, and have an evacuation plan for her child(ren) in case of disaster.
<sup>b</sup>Answered yes to having copies of important documents in a safe place outside of the home.
<sup>c</sup>Answered yes to having emergency supplies in the home for at least seven days and/or have emergency supplies to take with if she had to leave quickly.

Among 20.7% (n=184) of women reporting zero preparedness behaviors, significant differences were observed in urban or rural residence and race. Twenty-three percent of women living in an urban area and 16% of women living in a rural area reported zero preparedness behaviors (p=0.019). Of the six races reported, Japanese women had the highest proportion reporting zero preparedness behaviors, while Other Pacific Islanders had the lowest proportion reporting zero preparedness behaviors. Reporting of zero preparedness behaviors did not vary by education, marital status, insurance, family size, income, or WIC participation. (Table 4)

**Table 4.**
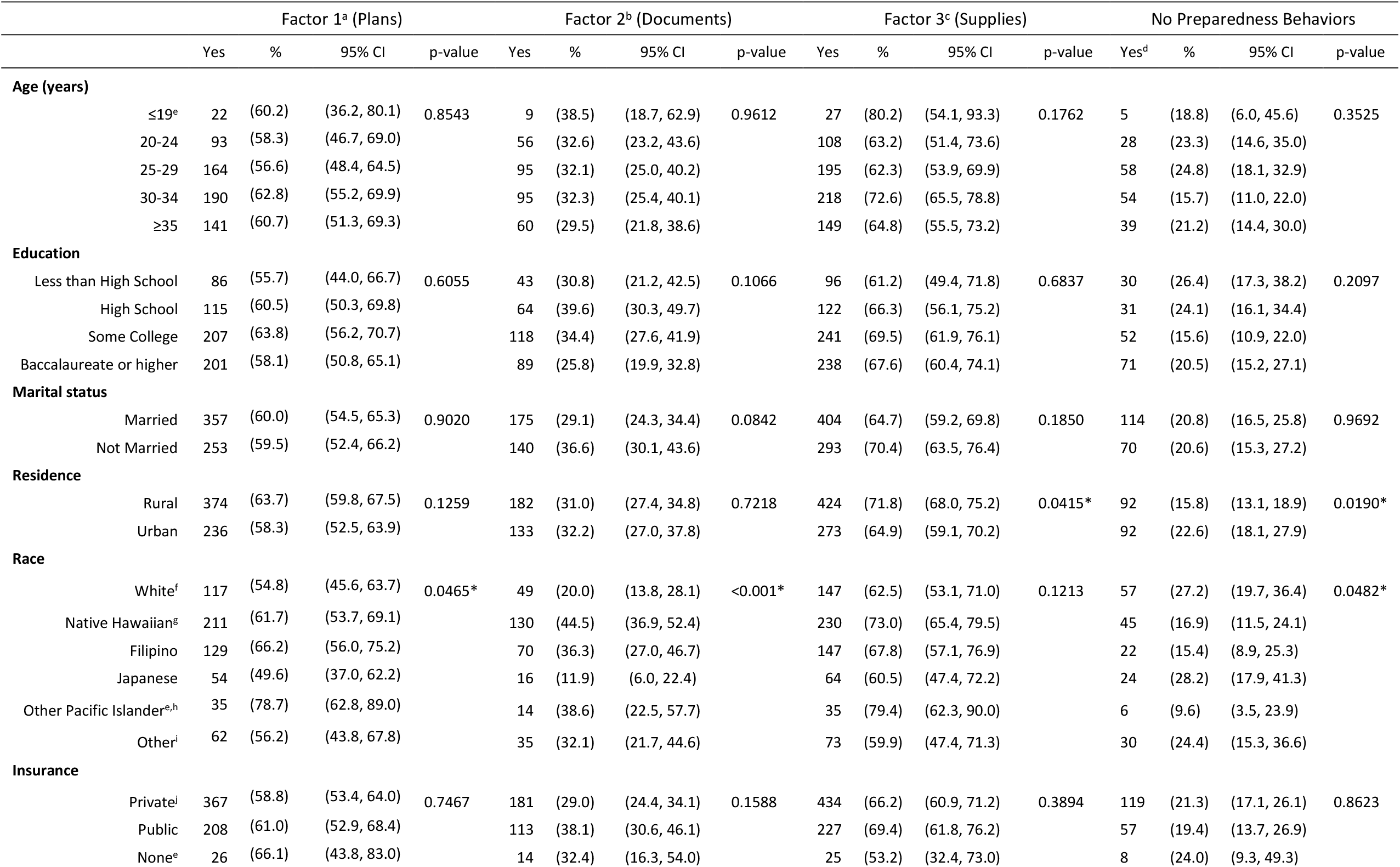
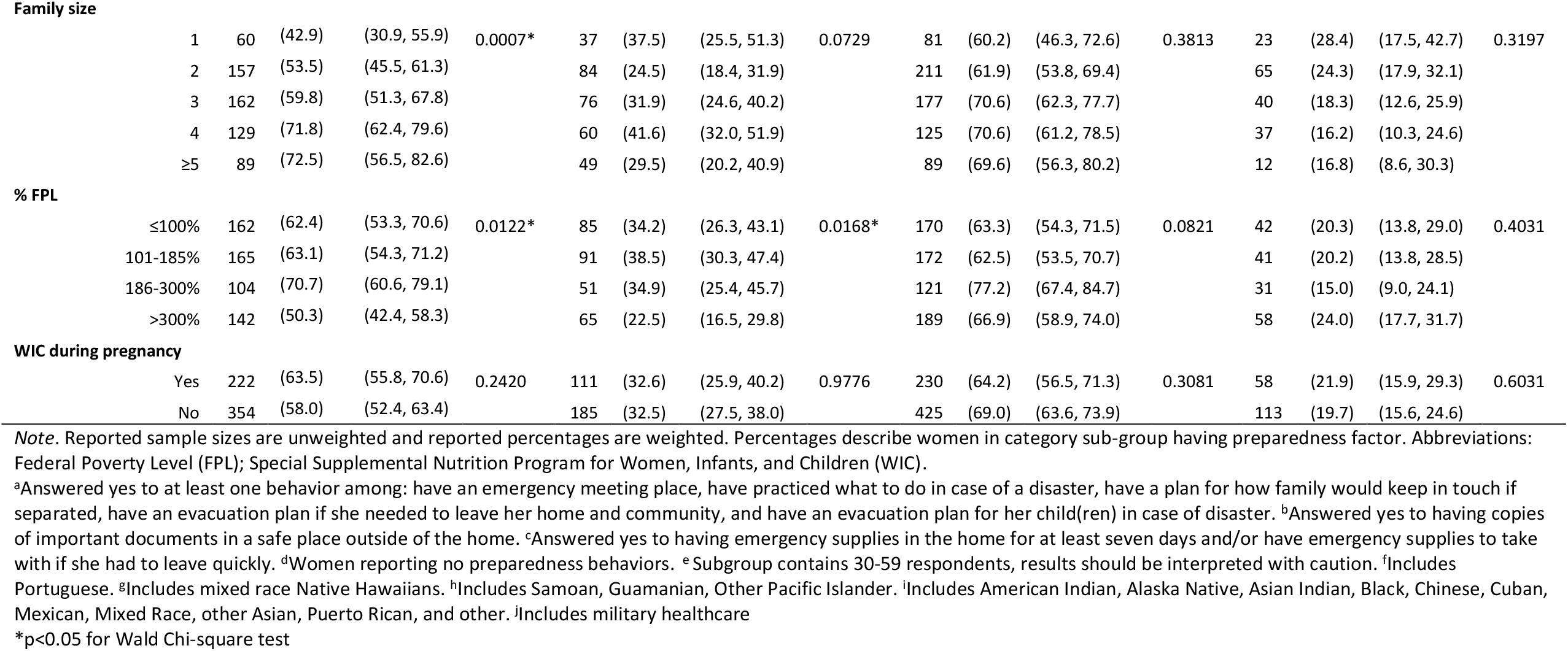
Emergency preparedness behaviors by demographic category among women with a recent live birth in Hawaii, 2016 Hawaii PRAMS

|  | Factor 1<sup>a</sup> (Plans) |  |  |  | Factor 2<sup>b</sup> (Documents) |  |  |  | Factor 3<sup>c</sup> (Supplies) |  |  |  | No Preparedness Behaviors |  |  |  |
| --- | --- | --- | --- | --- | --- | --- | --- | --- | --- | --- | --- | --- | --- | --- | --- | --- |
|  | Yes | % | 95% CI | p-value | Yes | % | 95% CI | p-value | Yes | % | 95% CI | p-value | Yes<sup>d</sup> | % | 95% CI | p-value |
| **Age (years)** |  |  |  |  |  |  |  |  |  |  |  |  |  |  |  |  |
| ≤19<sup>e</sup> | 22 | (60.2) | (36.2, 80.1) | 0.8543 | 9 | (38.5) | (18.7, 62.9) | 0.9612 | 27 | (80.2) | (54.1, 93.3) | 0.1762 | 5 | (18.8) | (6.0, 45.6) | 0.3525 |
| 20-24 | 93 | (58.3) | (46.7, 69.0) |  | 56 | (32.6) | (23.2, 43.6) |  | 108 | (63.2) | (51.4, 73.6) |  | 28 | (23.3) | (14.6, 35.0) |  |
| 25-29 | 164 | (56.6) | (48.4, 64.5) |  | 95 | (32.1) | (25.0, 40.2) |  | 195 | (62.3) | (53.9, 69.9) |  | 58 | (24.8) | (18.1, 32.9) |  |
| 30-34 | 190 | (62.8) | (55.2, 69.9) |  | 95 | (32.3) | (25.4, 40.1) |  | 218 | (72.6) | (65.5, 78.8) |  | 54 | (15.7) | (11.0, 22.0) |  |
| ≥35 | 141 | (60.7) | (51.3, 69.3) |  | 60 | (29.5) | (21.8, 38.6) |  | 149 | (64.8) | (55.5, 73.2) |  | 39 | (21.2) | (14.4, 30.0) |  |
| **Education** |  |  |  |  |  |  |  |  |  |  |  |  |  |  |  |  |
| Less than High School | 86 | (55.7) | (44.0, 66.7) | 0.6055 | 43 | (30.8) | (21.2, 42.5) | 0.1066 | 96 | (61.2) | (49.4, 71.8) | 0.6837 | 30 | (26.4) | (17.3, 38.2) | 0.2097 |
| High School | 115 | (60.5) | (50.3, 69.8) |  | 64 | (39.6) | (30.3, 49.7) |  | 122 | (66.3) | (56.1, 75.2) |  | 31 | (24.1) | (16.1, 34.4) |  |
| Some College | 207 | (63.8) | (56.2, 70.7) |  | 118 | (34.4) | (27.6, 41.9) |  | 241 | (69.5) | (61.9, 76.1) |  | 52 | (15.6) | (10.9, 22.0) |  |
| Baccalaureate or higher | 201 | (58.1) | (50.8, 65.1) |  | 89 | (25.8) | (19.9, 32.8) |  | 238 | (67.6) | (60.4, 74.1) |  | 71 | (20.5) | (15.2, 27.1) |  |
| **Marital status** |  |  |  |  |  |  |  |  |  |  |  |  |  |  |  |  |
| Married | 357 | (60.0) | (54.5, 65.3) | 0.9020 | 175 | (29.1) | (24.3, 34.4) | 0.0842 | 404 | (64.7) | (59.2, 69.8) | 0.1850 | 114 | (20.8) | (16.5, 25.8) | 0.9692 |
| Not Married | 253 | (59.5) | (52.4, 66.2) |  | 140 | (36.6) | (30.1, 43.6) |  | 293 | (70.4) | (63.5, 76.4) |  | 70 | (20.6) | (15.3, 27.2) |  |
| **Residence** |  |  |  |  |  |  |  |  |  |  |  |  |  |  |  |  |
| Rural | 374 | (63.7) | (59.8, 67.5) | 0.1259 | 182 | (31.0) | (27.4, 34.8) | 0.7218 | 424 | (71.8) | (68.0, 75.2) | 0.0415* | 92 | (15.8) | (13.1, 18.9) | 0.0190* |
| Urban | 236 | (58.3) | (52.5, 63.9) |  | 133 | (32.2) | (27.0, 37.8) |  | 273 | (64.9) | (59.1, 70.2) |  | 92 | (22.6) | (18.1, 27.9) |  |
| **Race** |  |  |  |  |  |  |  |  |  |  |  |  |  |  |  |  |
| White<sup>f</sup> | 117 | (54.8) | (45.6, 63.7) | 0.0465* | 49 | (20.0) | (13.8, 28.1) | <0.001* | 147 | (62.5) | (53.1, 71.0) | 0.1213 | 57 | (27.2) | (19.7, 36.4) | 0.0482* |
| Native Hawaiian<sup>g</sup> | 211 | (61.7) | (53.7, 69.1) |  | 130 | (44.5) | (36.9, 52.4) |  | 230 | (73.0) | (65.4, 79.5) |  | 45 | (16.9) | (11.5, 24.1) |  |
| Filipino | 129 | (66.2) | (56.0, 75.2) |  | 70 | (36.3) | (27.0, 46.7) |  | 147 | (67.8) | (57.1, 76.9) |  | 22 | (15.4) | (8.9, 25.3) |  |
| Japanese | 54 | (49.6) | (37.0, 62.2) |  | 16 | (11.9) | (6.0, 22.4) |  | 64 | (60.5) | (47.4, 72.2) |  | 24 | (28.2) | (17.9, 41.3) |  |
| Other Pacific Islander<sup>e,h</sup> | 35 | (78.7) | (62.8, 89.0) |  | 14 | (38.6) | (22.5, 57.7) |  | 35 | (79.4) | (62.3, 90.0) |  | 6 | (9.6) | (3.5, 23.9) |  |
| Other<sup>i</sup> | 62 | (56.2) | (43.8, 67.8) |  | 35 | (32.1) | (21.7, 44.6) |  | 73 | (59.9) | (47.4, 71.3) |  | 30 | (24.4) | (15.3, 36.6) |  |
| **Insurance** |  |  |  |  |  |  |  |  |  |  |  |  |  |  |  |  |
| Private<sup>j</sup> | 367 | (58.8) | (53.4, 64.0) | 0.7467 | 181 | (29.0) | (24.4, 34.1) | 0.1588 | 434 | (66.2) | (60.9, 71.2) | 0.3894 | 119 | (21.3) | (17.1, 26.1) | 0.8623 |
| Public | 208 | (61.0) | (52.9, 68.4) |  | 113 | (38.1) | (30.6, 46.1) |  | 227 | (69.4) | (61.8, 76.2) |  | 57 | (19.4) | (13.7, 26.9) |  |
| None<sup>e</sup> | 26 | (66.1) | (43.8, 83.0) |  | 14 | (32.4) | (16.3, 54.0) |  | 25 | (53.2) | (32.4, 73.0) |  | 8 | (24.0) | (9.3, 49.3) |  |
| **Family size** |  |  |  |  |  |  |  |  |  |  |  |  |  |  |  |  |
| 1 | 60 | (42.9) | (30.9, 55.9) | 0.0007* | 37 | (37.5) | (25.5, 51.3) | 0.0729 | 81 | (60.2) | (46.3, 72.6) | 0.3813 | 23 | (28.4) | (17.5, 42.7) | 0.3197 |
| 2 | 157 | (53.5) | (45.5, 61.3) |  | 84 | (24.5) | (18.4, 31.9) |  | 211 | (61.9) | (53.8, 69.4) |  | 65 | (24.3) | (17.9, 32.1) |  |
| 3 | 162 | (59.8) | (51.3, 67.8) |  | 76 | (31.9) | (24.6, 40.2) |  | 177 | (70.6) | (62.3, 77.7) |  | 40 | (18.3) | (12.6, 25.9) |  |
| 4 | 129 | (71.8) | (62.4, 79.6) |  | 60 | (41.6) | (32.0, 51.9) |  | 125 | (70.6) | (61.2, 78.5) |  | 37 | (16.2) | (10.3, 24.6) |  |
| ≥5 | 89 | (72.5) | (56.5, 82.6) |  | 49 | (29.5) | (20.2, 40.9) |  | 89 | (69.6) | (56.3, 80.2) |  | 12 | (16.8) | (8.6, 30.3) |  |
| **% FPL** |  |  |  |  |  |  |  |  |  |  |  |  |  |  |  |  |
| ≤100% | 162 | (62.4) | (53.3, 70.6) | 0.0122* | 85 | (34.2) | (26.3, 43.1) | 0.0168* | 170 | (63.3) | (54.3, 71.5) | 0.0821 | 42 | (20.3) | (13.8, 29.0) | 0.4031 |
| 101-185% | 165 | (63.1) | (54.3, 71.2) |  | 91 | (38.5) | (30.3, 47.4) |  | 172 | (62.5) | (53.5, 70.7) |  | 41 | (20.2) | (13.8, 28.5) |  |
| 186-300% | 104 | (70.7) | (60.6, 79.1) |  | 51 | (34.9) | (25.4, 45.7) |  | 121 | (77.2) | (67.4, 84.7) |  | 31 | (15.0) | (9.0, 24.1) |  |
| >300% | 142 | (50.3) | (42.4, 58.3) |  | 65 | (22.5) | (16.5, 29.8) |  | 189 | (66.9) | (58.9, 74.0) |  | 58 | (24.0) | (17.7, 31.7) |  |
| **WIC during pregnancy** |  |  |  |  |  |  |  |  |  |  |  |  |  |  |  |  |
| Yes | 222 | (63.5) | (55.8, 70.6) | 0.2420 | 111 | (32.6) | (25.9, 40.2) | 0.9776 | 230 | (64.2) | (56.5, 71.3) | 0.3081 | 58 | (21.9) | (15.9, 29.3) | 0.6031 |
| No | 354 | (58.0) | (52.4, 63.4) |  | 185 | (32.5) | (27.5, 38.0) |  | 425 | (69.0) | (63.6, 73.9) |  | 113 | (19.7) | (15.6, 24.6) |  |

*Note.* Reported sample sizes are unweighted and reported percentages are weighted. Percentages describe women in category sub-group having preparedness factor. Abbreviations: Federal Poverty Level (FPL); Special Supplemental Nutrition Program for Women, Infants, and Children (WIC).
<sup>a</sup>Answered yes to at least one behavior among: have an emergency meeting place, have practiced what to do in case of a disaster, have a plan for how family would keep in touch if separated, have an evacuation plan if she needed to leave her home and community, and have an evacuation plan for her child(ren) in case of disaster. <sup>b</sup>Answered yes to having copies of important documents in a safe place outside of the home. <sup>c</sup>Answered yes to having emergency supplies in the home for at least seven days and/or have emergency supplies to take with if she had to leave quickly. <sup>d</sup>Women reporting no preparedness behaviors. <sup>e</sup>Subgroup contains 30-59 respondents, results should be interpreted with caution. <sup>f</sup>Includes Portuguese. <sup>g</sup>Includes mixed race Native Hawaiians. <sup>h</sup>Includes Samoan, Guamanian, Other Pacific Islander. <sup>i</sup>Includes American Indian, Alaska Native, Asian Indian, Black, Chinese, Cuban, Mexican, Mixed Race, other Asian, Puerto Rican, and other. <sup>j</sup>Includes military healthcare
\*p<0.05 for Wald Chi-square test

We assessed factor loading plots and determined the eight preparedness behaviors could be described by three factors (Online Resources 1-3). The first factor, having emergency plans, captured women that responded yes to at least one of the behaviors: emergency meeting place, practiced what to do, plan to keep in touch, plan for themselves to evacuate, or an evacuation plan for their child(ren). The second factor included women who responded yes to having copies of important documents, and the third factor, included women that had emergency supplies for at least seven days and/or had emergency supplies prepared if they had to leave quickly.

Within the sample, 59.8% (95% CI: 55.5-64.0%) of women had emergency plans; 31.8% (95% CI: 27.9-36.0%) had copies of important documents; and 66.8% (95% CI: 62.5-70.8%) had emergency supplies (Table 3). Having emergency plans varied by race; among Other Pacific Islanders, 79% reported an emergency planning behavior compared to 50% of Japanese women. Emergency planning also varied by family size and percent of the FPL. More than 70% of women with a family size of four or greater reported having emergency plans, while 53.5% (95% CI: 45.5, 61.3) of women with a family size of two reported these behaviors. Among women with an income of 186-300% FPL, 70.7% (95% CI: 60.6, 79.1) reported having emergency plans, whereas 50.3% (95% CI: 60.6, 79.1) of women with an income more than 300% FPL reported emergency planning behaviors. Demographic characteristics among women reporting having copies of important documents varied significantly by race and FPL. Having copies of important documents was most commonly reported among Native Hawaiians (44.5%, 95% CI: 36.9, 52.4). In contrast, 11.9% (95% CI: 6.0, 22.4) of Japanese women reported having copies of important documents. A greater percent of women living at 101-185% FPL (38.5%, 95% CI: 30.3, 47.4) had copies of important documents compared to 22.5% (95% CI: 16.5, 29.8) of women living at more than 300% FPL. Reporting of emergency supplies varied by urban or rural residence. Sixteen percent of women living in a rural residence (15.8%, 95% CI: 13.1, 18.9) and 23.6% (95% CI: 18.1, 27.9) of women living in an urban residence had emergency supplies. (Table 4)

### Models

The results of the multivariable analysis are presented in Table 5. Results from women 19-years-old and younger, Other Pacific Islanders, and those with no insurance at the time of survey, should be interpreted with caution as their sample size was small (30-59 respondents). Education and marital status were not associated with any preparedness factors. The prevalence of completing at least one planning emergency preparedness behavior differed significantly by race, family size, and poverty level. Other Pacific Islanders were 45% more likely to report having emergency plans compared to White women (aPR 1.45, 95% CI: 1.09-1.93). Women with a family size of one were 33% (aPR 0.67, 95% CI: 0.46-0.96) less likely to report having emergency plans compared to women with a family size of two. In contrast, having emergency supplies is more common among women with a family size of four (aPR 1.31, 95% CI: 1.06, 1.61) and five or more (aPR 1.33, 95% CI: 1.04-1.70) compared to those with a family size of two. Compared to women with an income above 300% FPL, those with an income between 186 and 300% FPL were 28% (aPR 1.28, 95% CI: 1.03-1.60) more likely to have emergency plans.

**Table 5.** Multivariable logistic regression of the characteristics of women with a recent live birth in Hawaii on the presence of emergency preparedness behavior factors, 2016 Hawaii PRAMS

|  |  | Factor 1<sup>a</sup> (Plans) |  | Factor 2<sup>b</sup> (Documents) |  | Factor 3<sup>c</sup> (Supplies) |  |
| --- | --- | --- | --- | --- | --- | --- | --- |
|  |  | Prevalence Ratio | 95% CI | Prevalence Ratio | 95% CI | Prevalence Ratio | 95% CI |
| **Age (years)** |  |  |  |  |  |  |  |
|  | ≤19<sup>d</sup> | 1.27 | (0.96, 1.69) | 1.33 | (0.74, 2.42) | 1.33 | (1.14, 1.55) |
|  | 20-24 | 1.06 | (0.82, 1.38) | 1.21 | (0.80, 1.84) | 1.00 | (0.81, 1.25) |
|  | 25-29 | 0.96 | (0.79, 1.18) | 1.01 | (0.71, 1.45) | 0.92 | (0.77, 1.09) |
|  | 30-34 | Referent |  | Referent |  | Referent |  |
|  | ≥35 | 1.00 | (0.82, 1.23) | 1.08 | (0.75, 1.56) | 0.90 | (0.75, 1.09) |
| **Education** |  |  |  |  |  |  |  |
|  | Less than High School | 0.81 | (0.59, 1.11) | 0.76 | (0.45, 1.30) | 0.89 | (0.69, 1.14) |
|  | High School | 0.95 | (0.75, 1.21) | 1.25 | (0.85, 1.84) | 0.93 | (0.75, 1.16) |
|  | Some College | 1.02 | (0.85, 1.23) | 1.06 | (0.75, 1.51) | 1.01 | (0.86, 1.17) |
|  | Baccalaureate or higher | Referent |  | Referent |  | Referent |  |
| **Marital status** |  |  |  |  |  |  |  |
|  | Married | Referent |  | Referent |  | Referent |  |
|  | Not Married | 0.98 | (0.80, 1.19) | 0.89 | (0.64, 1.25) | 1.14 | (0.98, 1.33) |
| **Residence** |  |  |  |  |  |  |  |
|  | Rural | 1.08 | (0.95, 1.23) | 0.75 | (0.60, 0.95) | 1.10 | (0.99, 1.23) |
|  | Urban | Referent |  | Referent |  | Referent |  |
| **Race** |  |  |  |  |  |  |  |
|  | White<sup>e</sup> | Referent |  | Referent |  | Referent |  |
|  | Native Hawaiian<sup>f</sup> | 1.16 | (0.93, 1.44) | 2.16 | (1.45, 3.24) | 1.18 | (0.98, 1.42) |
|  | Filipino | 1.23 | (0.98, 1.55) | 1.69 | (1.06, 2.69) | 1.07 | (0.87, 1.32) |
|  | Japanese | 1.06 | (0.78, 1.43) | 0.75 | (0.34, 1.69) | 0.99 | (0.76, 1.29) |
|  | Other Pacific Islander<sup>d,g</sup> | 1.45 | (1.09, 1.93) | 2.02 | (1.10, 3.71) | 1.44 | (1.18, 1.76) |
|  | Other<sup>h</sup> | 1.03 | (0.77, 1.38) | 1.50 | (0.89, 2.52) | 1.02 | (0.80, 1.30) |
| **Insurance** |  |  |  |  |  |  |  |
|  | Private<sup>i</sup> | Referent |  | Referent |  | Referent |  |
|  | Public | 1.02 | (0.85, 1.23) | 1.42 | (1.04, 1.93) | 1.06 | (0.91, 1.24) |
|  | None<sup>d</sup> | 0.91 | (0.60, 1.38) | 0.92 | (0.38, 2.27) | 0.81 | (0.53, 1.25) |
| **Family size** |  |  |  |  |  |  |  |
|  | 1 | 0.67 | (0.46, 0.96) | 1.26 | (0.80, 1.97) | 0.86 | (0.64, 1.15) |
|  | 2 | Referent |  | Referent |  | Referent |  |
|  | 3 | 1.09 | (0.88, 1.35) | 1.23 | (0.85, 1.78) | 1.12 | (0.94, 1.34) |
|  | 4 | 1.31 | (1.06, 1.61) | 1.50 | (1.02, 2.20) | 1.19 | (0.99, 1.43) |
|  | ≥5 | 1.33 | (1.04, 1.70) | 1.02 | (0.62, 1.67) | 1.24 | (1.02, 1.52) |
| **% FPL** |  |  |  |  |  |  |  |
|  | ≤100% | 1.05 | (0.77, 1.43) | 1.07 | (0.65, 1.75) | 0.74 | (0.57, 0.97) |
|  | 101-185% | 1.01 | (0.79, 1.31) | 1.47 | (0.99, 2.20) | 0.84 | (0.69, 1.02) |
|  | 186-300% | 1.28 | (1.03, 1.60) | 1.41 | (0.94, 2.10) | 1.10 | (0.94, 1.28) |
|  | >300% | Referent |  | Referent |  | Referent |  |
| **WIC during pregnancy** |  |  |  |  |  |  |  |
|  | Yes | 1.11 | (0.94, 1.31) | 0.72 | (0.53, 0.98) | 0.96 | (0.82, 1.12) |
|  | No | Referent |  | Referent |  | Referent |  |

*Note.* Reported prevalence ratios are adjusted for all variables included in models. Abbreviations: Federal Poverty Level (FPL); Special Supplemental Nutrition Program for Women, Infants, and Children (WIC).
<sup>a</sup>Answered yes to at least one behavior among: have an emergency meeting place, have practiced what to do in case of a disaster, have a plan for how family would keep in touch if separated, have an evacuation plan if she needed to leave her home and community, and have an evacuation plan for her child(ren) in case of disaster. <sup>b</sup>Answered yes to having copies of important documents in a safe place outside of the home. <sup>c</sup>Answered yes to having emergency supplies in the home for at least seven days and/or have emergency supplies to take with if she had to leave quickly.
<sup>d</sup>Subgroup contains 30-59 respondents, results should be interpreted with caution. <sup>e</sup>Includes Portuguese. <sup>f</sup>Includes mixed race Native Hawaiians.
<sup>g</sup>Includes Samoan, Guamanian, Other Pacific Islander. <sup>h</sup>Includes American Indian, Alaska Native, Asian Indian, Black, Chinese, Cuban, Mexican, Mixed Race, other Asian, Puerto Rican, and other. <sup>i</sup>Includes military healthcare.

Having copies of important documents significantly differed in the adjusted models by residence, race, insurance status, family size, and WIC participation. This behavior was less common among women living in a rural residence compared to women living in an urban residence (aPR 0.75, 95% CI: 0.60, 0.95), and among women that used WIC during pregnancy compared to those that did not (aPR 0.72, 95% CI: 0.53, 0.98). Native Hawaiian (aPR 2.16, 95% CI: 1.45-3.24), Filipino (aPR 1.69, 95% CI:1.06-2.69), and Other Pacific Islanders (aPR 2.02, 95% CI: 1.10-3.71) were more likely than White women to have copies of important documents. Additionally, having copies of important documents was more common among women with public insurance compared to women with private insurance (aPR 1.42, 95% CI: 1.04, 1.93), and among women with a family size of four compared to women with a family size of two (aPR 1.50, 95% CI: 1.02, 2.20).

Significant differences were noted by age, race, family size, and FPL in the adjusted model for emergency supplies. Women 19-years-old and younger were 33% more likely to have emergency supplies compared women 30-34 years old (aPR 1.33, 95% CI: 1.14-1.55). Having emergency supplies was also more likely among Other Pacific Islanders compared to White women (aPR 1.44, 95% CI: 1.18-1.76), and women with a family size of five or more, compared to those with a family size of two. Women with an income at or less than 100% FPL were 26% (aPR 0.74, 95% CI: 0.57-0.97) less likely to have emergency supplies compared to women with an income more than 300% FPL.

## DISCUSSION

Analysis of the 2016 Hawaii PRAMS data shows the eight preparedness behaviors can be generalized into three factors - having emergency plans, having copies of important documents, and having emergency supplies. About 80% of women participated in at least one preparedness behavior, and each behavior displayed at least 30% participation.

Race and ethnicity have previously been shown to be important predictors of preparedness, although no consensus exists between the direction of association of race and preparedness in current literature (Zilversmit, Sappenfield, Zotti, and McGehee 2014; DeBastiani, Strine, Vagi, Barnett, and Kahn 2015; Baker 2011; Murphy, Cody, Frank, Glik, and Ang 2009; Diekman, Kearney, O’Neil, and Mack 2007; Bethel, Burke, and Britt 2013). An assessment of the 2006 - 2010 Behavioral Risk Factor Surveillance System general preparedness module found Black and Hispanic respondents were more likely to have a three-day supply of water, and an evacuation plan prepared compared to White respondents; however White respondents were more likely to have a three-day supply of food, battery operated radio, and medication (Bethel, Burke, and Britt 2013). Among a nationally representative sample of US households completing an online survey to assess predictors of disaster preparedness and compliance, the authors found that non-White individuals were more likely to have emergency plans (Murphy, Cody, Frank, Glik, and Ang 2009). In our analysis, Other Pacific Islanders reported higher participation in all three factors than White women. Although the sample size of Other Pacific Islanders was small, PRAMS methodology is designed to be representative of the state. Other Pacific Islanders are a minority group in the US, however many reside within Hawaii, making Hawaii the ideal state to assess behaviors among Other Pacific Islander women with a recent live birth (Office of Minority Health 2020).

Family size was significantly associated with each factor in multivariable analyses. A family size of one was associated with a lower likelihood of having emergency plans compared to a family size of two. Compared to women with a family size of two, family sizes of four or more were associated with increased likelihood of having emergency plans, a family size of four was associated with a higher likelihood of having copies of important documents, and a family size of five or more was associated with a higher likelihood of having emergency supplies. A study by Zilversmit et al. (2014) observed families with five or more members were 30% more likely to have an emergency plan compared to families of one to four members when assessing the presence of an emergency plan among postpartum women in Arkansas. A focus group discussing household emergency preparedness among homeowners found children in the home promote preparedness for two reasons: it is a way for parents to protect their children, and preparedness is a result of increased involvement in community activities that prompt preparedness behaviors (Diekman, Kearney, O’Neil, and Mack 2007).

In this study, income categorized by percent of the FPL was significant in the emergency plans and emergency supplies adjusted models. Women with an income at or below 100% FPL were less likely to have emergency supplies compared to women with an income greater than 300% FPL, but there were no differences between these groups for emergency plans. Women with an income 186%-300 FPL were more likely to have emergency plans than women with an income greater than 300% FPL. Data from the 2016 Styles survey suggests among US adults, cost is a barrier to emergency preparedness, while discounts to buy preparedness supplies are a motivator for emergency preparedness (Kruger et al. 2020). Buying surplus supplies in case of an emergency may be economically burdensome.

The second factor, having copies of important documents, was associated with residence, current insurance status, and participation in WIC. Women living in rural areas of Hawaii were less likely to report having copies of important documents in a safe location outside of the home compared to women with an urban residence. A rural area may have fewer options for safe storage of important documents outside of the home. Having copies of important documents was 42% more likely among women with public insurance, compared to women with private insurance. Use of WIC was associated with a lower prevalence of having copies of important documents, compared to those not using WIC. Participation in WIC during pregnancy requires a woman to be classified as having a nutritional risk by a health professional, and have an income at or below 185% FPL (USDA Food and Nutrition Service). Although percent FPL was not a significant predictor of having copies of important documents in this study, the income thresholds for WIC eligibility may be related to its association with lower prevalence of having copies of important documents.

In this analysis, younger age was significantly associated with having emergency supplies. In contrast, a study among Florida residents identified those aged 40-70 were more prepared than other respondents (Baker 2011).

Our study identified minimal demographic differences with preparedness behaviors among women with a recent live birth in Hawaii, suggesting disaster preparedness interventions should target all pregnant and postpartum women. Among mailed surveys, shorter questionnaires are associated with higher response rates (Edwards et al. 2009; Yammarino, Skinner, and Childers 1991). Therefore, reducing this eight-part preparedness question to three-parts identified through factor analysis may be preferable. This study is limited by self-reported data so misclassification and reporting bias may be present. Furthermore, dichotomous answer options do not capture levels of preparedness, nor specify a timeframe to consider when responding. This analysis is limited as Hawaii PRAMS does not collect information on social support and nontangible resources; however, these have been described as a strong predictor of preparedness (Diekman, Kearney, O’Neil, and Mack 2007; Kim, Pant, and Yamashita 2018; Levac, Toal-Sullivan, and O’Sullivan 2012; Giarratano, Barcelona, Savage, and Harville 2019; Ehrlich et al. 2010).

## CONCLUSION

This study provides a measure of emergency preparedness among women with a recent live birth in Hawaii and is the first to describe a methodology to analyze the eight-part PRAMS emergency preparedness question. If other states observe similar factor analysis results of these eight preparedness behaviors, this PRAMS question could be reformatted to three parts. Fewer questions may increase the use of this question by jurisdictions and participant response rates may improve. Additionally, these results can inform Hawaii’s efforts to increasing disaster preparedness among postpartum women and families in Hawaii. The effect of interventions to increase preparedness can be measured by tracking this question over time.

## Supporting information

Supplemental Resources

## Declarations

### Ethics approval

PRAMS protocol has been reviewed and approved by Centers for Disease Control and Prevention’s Institutional Review Board and approved as human subjects research (HSR #2233)

### Consent to participate

All survey respondents provided verbal consent via phone or implied consent by returning a completed questionnaire.

### Consent for publication

All authors have consented for publication.

### Availability of data and material

Data are available through request at https://www.cdc.gov/prams/prams-data/researchers.htm

### Code availability

SAS-callable SUDAAN code available upon request.

### Authors’ contributions

All authors have reviewed, contributed significantly to, and approve the paper.

### Data Availability

Availability of data and material Data are available through request at https://www.cdc.gov/prams/prams-data/researchers.htm

---

STROBE Statement—Checklist of items that should be included in reports of ***cross-sectional studies***

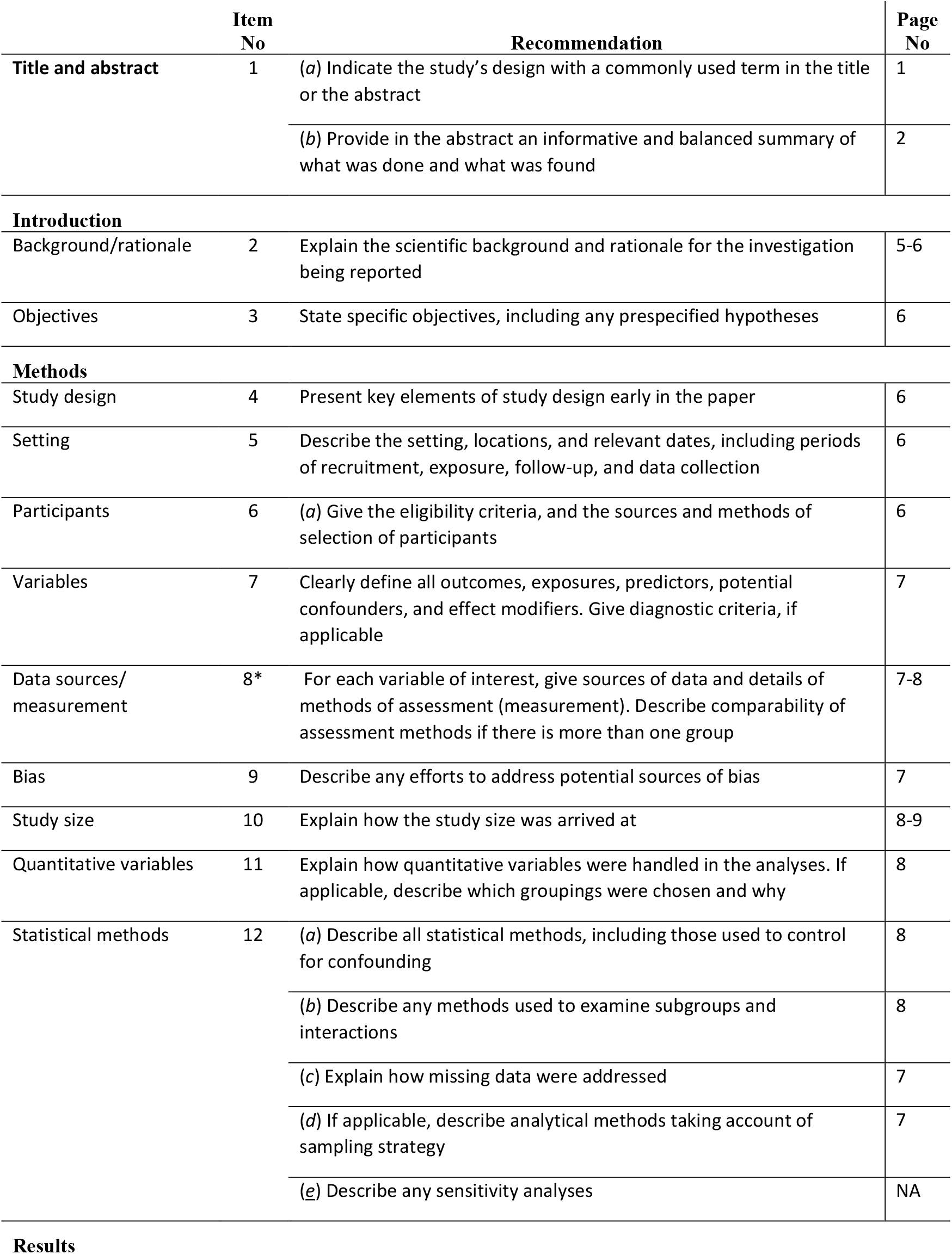
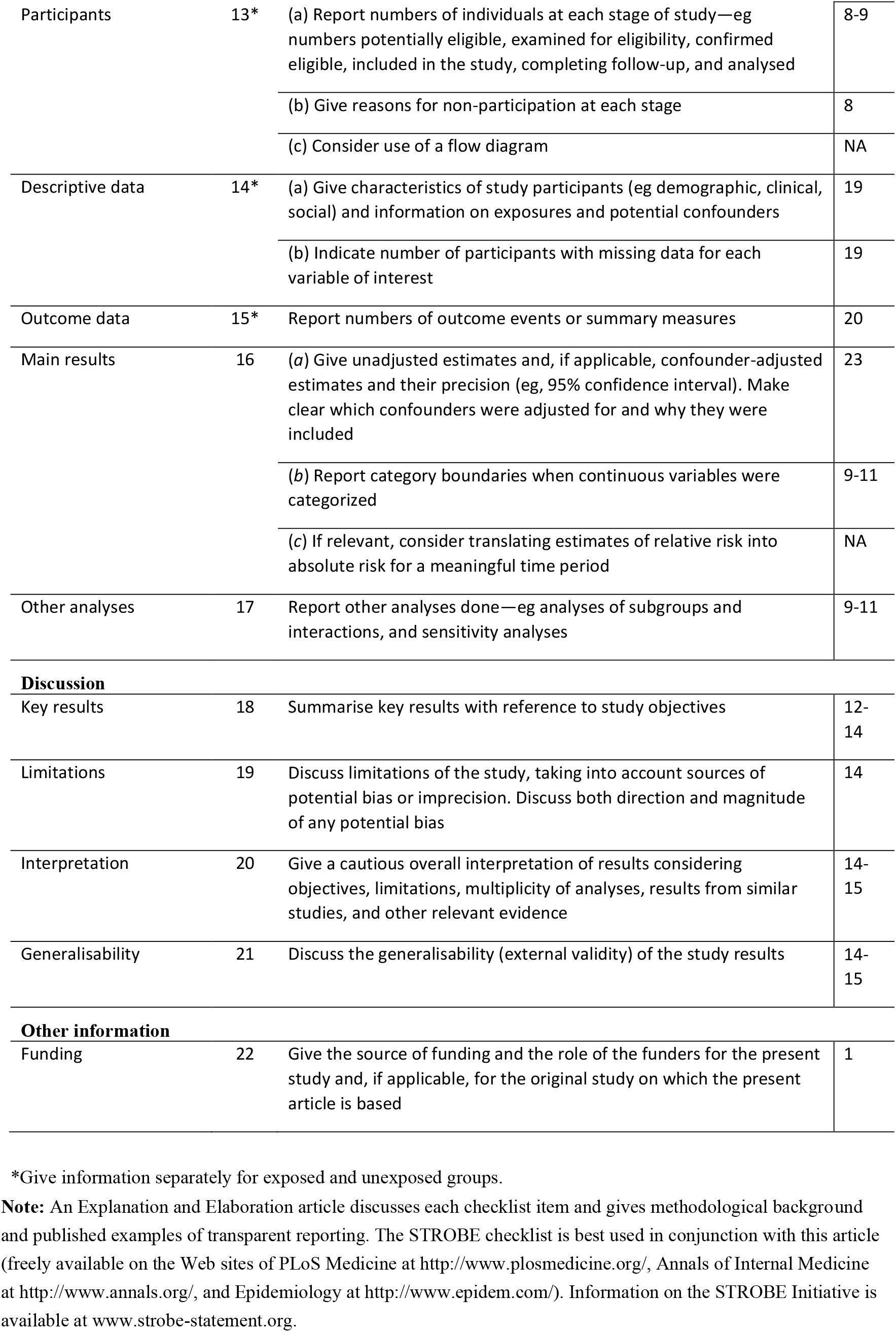

|  | Item No | Recommendation | Page No |
| --- | --- | --- | --- |
| **Title and abstract** | 1 | (a) Indicate the study's design with a commonly used term in the title or the abstract | 1 |
|  |  | (b) Provide in the abstract an informative and balanced summary of what was done and what was found | 2 |
| **Introduction** |  |  |  |
| Background/rationale | 2 | Explain the scientific background and rationale for the investigation being reported | 5-6 |
| Objectives | 3 | State specific objectives, including any prespecified hypotheses | 6 |
| **Methods** |  |  |  |
| Study design | 4 | Present key elements of study design early in the paper | 6 |
| Setting | 5 | Describe the setting, locations, and relevant dates, including periods of recruitment, exposure, follow-up, and data collection | 6 |
| Participants | 6 | (a) Give the eligibility criteria, and the sources and methods of selection of participants | 6 |
| Variables | 7 | Clearly define all outcomes, exposures, predictors, potential confounders, and effect modifiers. Give diagnostic criteria, if applicable | 7 |
| Data sources/<br>measurement | 8* | For each variable of interest, give sources of data and details of methods of assessment (measurement). Describe comparability of assessment methods if there is more than one group | 7-8 |
| Bias | 9 | Describe any efforts to address potential sources of bias | 7 |
| Study size | 10 | Explain how the study size was arrived at | 8-9 |
| Quantitative variables | 11 | Explain how quantitative variables were handled in the analyses. If applicable, describe which groupings were chosen and why | 8 |
| Statistical methods | 12 | (a) Describe all statistical methods, including those used to control for confounding | 8 |
|  |  | (b) Describe any methods used to examine subgroups and interactions | 8 |
|  |  | (c) Explain how missing data were addressed | 7 |
|  |  | (d) If applicable, describe analytical methods taking account of sampling strategy | 7 |
|  |  | (e) Describe any sensitivity analyses | NA |
| Participants | 13* | (a) Report numbers of individuals at each stage of study—eg numbers potentially eligible, examined for eligibility, confirmed eligible, included in the study, completing follow-up, and analysed | 8-9 |
|  |  | (b) Give reasons for non-participation at each stage | 8 |
|  |  | (c) Consider use of a flow diagram | NA |
| Descriptive data | 14* | (a) Give characteristics of study participants (eg demographic, clinical, social) and information on exposures and potential confounders | 19 |
|  |  | (b) Indicate number of participants with missing data for each variable of interest | 19 |
| Outcome data | 15* | Report numbers of outcome events or summary measures | 20 |
| Main results | 16 | (a) Give unadjusted estimates and, if applicable, confounder-adjusted estimates and their precision (eg, 95% confidence interval). Make clear which confounders were adjusted for and why they were included | 23 |
|  |  | (b) Report category boundaries when continuous variables were categorized | 9-11 |
|  |  | (c) If relevant, consider translating estimates of relative risk into absolute risk for a meaningful time period | NA |
| Other analyses | 17 | Report other analyses done—eg analyses of subgroups and interactions, and sensitivity analyses | 9-11 |
| **Discussion** |  |  |  |
| Key results | 18 | Summarise key results with reference to study objectives | 12-14 |
| Limitations | 19 | Discuss limitations of the study, taking into account sources of potential bias or imprecision. Discuss both direction and magnitude of any potential bias | 14 |
| Interpretation | 20 | Give a cautious overall interpretation of results considering objectives, limitations, multiplicity of analyses, results from similar studies, and other relevant evidence | 14-15 |
| Generalisability | 21 | Discuss the generalisability (external validity) of the study results | 14-15 |
| **Other information** |  |  |  |
| Funding | 22 | Give the source of funding and the role of the funders for the present study and, if applicable, for the original study on which the present article is based | 1 |

\*Give information separately for exposed and unexposed groups.
**Note:** An Explanation and Elaboration article discusses each checklist item and gives methodological background and published examples of transparent reporting. The STROBE checklist is best used in conjunction with this article (freely available on the Web sites of PLoS Medicine at http://www.plosmedicine.org/, Annals of Internal Medicine at http://www.annals.org/, and Epidemiology at http://www.epidem.com/). Information on the STROBE Initiative is available at www.strobe-statement.org.

