## Supplemental Resources for "Disaster preparedness among women with a recent live birth in Hawaii, a cross-sectional study, results from the Pregnancy Risk Assessment Monitoring System (PRAMS), 2016"

**Supplementary Information**

**Online Resources 1-3** Factor loading plots resulting from maximum likelihood factor analysis with varimax rotation. A, I have an emergency meeting place for family members (other than my home); B, My family and I have practiced what to do in case of a disaster; C, I have a plan for how my family and I would keep in touch if we were separated; D, I have an evacuation plan if I need to leave my home and community; E, I have an evacuation plan for my child or children in case of disaster (permission for day care or school to release my child to another adult); F, I have copies of important documents like birth certificates and insurance policies in a safe place outside of my home; G, I have emergency supplies in my home for my family such as enough extra water, food, and medicine to last for at least seven days; H, I have emergency supplies that I keep in my car, at work, or at home to take with me if I have to leave quickly.

**Online Resource 1** Factor loading plot between factor 1 (plans) and factor 2 (documents)

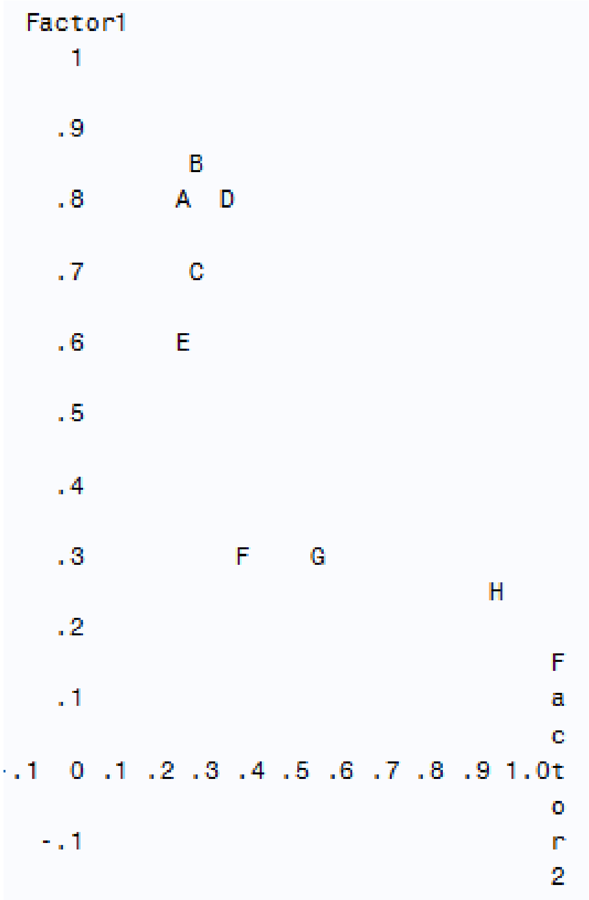

**Online Resource 2** Factor loading plot between factor 1 (plans) and factor 3 (supplies)

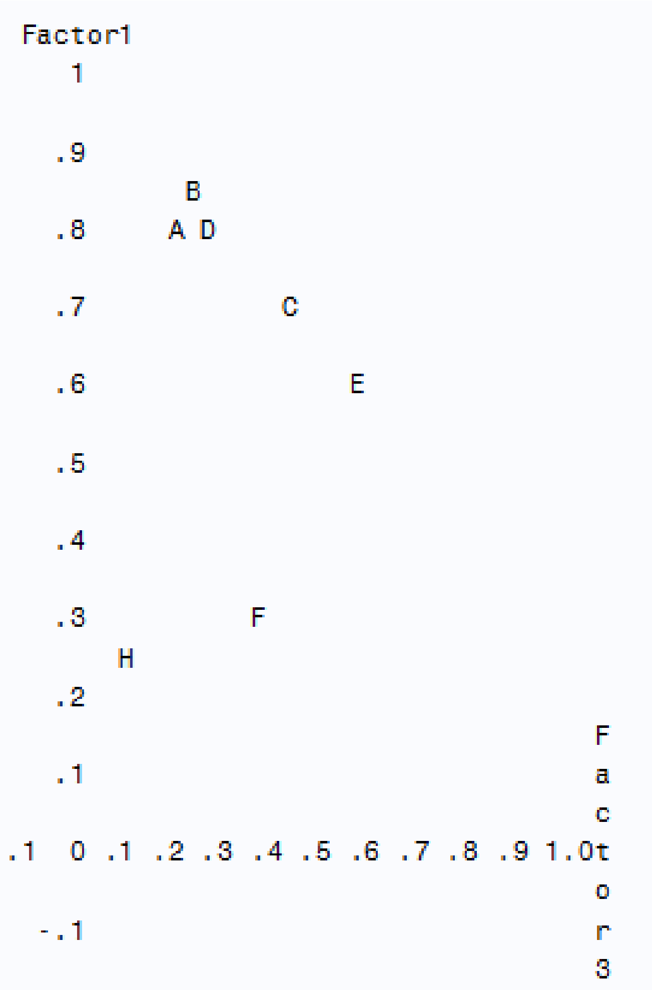

**Online Resource 3** Factor loading plot between factor 2 (documents) and factor 3 (supplies)

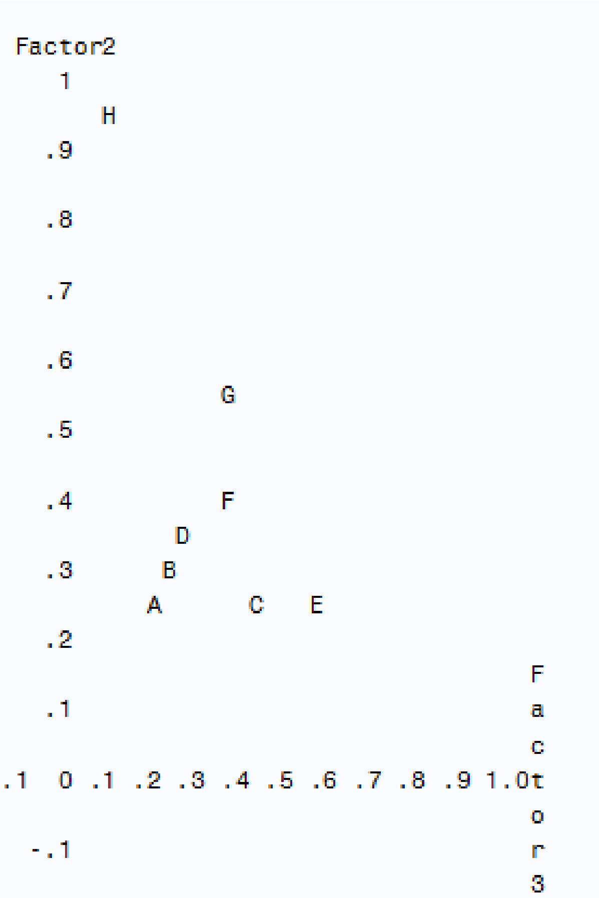
